# SARS-CoV-2 Distribution in Residential Housing Suggests Contact Deposition and Correlates with *Rothia* sp

**DOI:** 10.1101/2021.12.06.21267101

**Authors:** Victor J Cantú, Rodolfo A. Salido, Shi Huang, Gibraan Rahman, Rebecca Tsai, Holly Valentine, Celestine G. Magallanes, Stefan Aigner, Nathan A. Baer, Tom Barber, Pedro Belda-Ferre, Maryann Betty, MacKenzie Bryant, Martin Casas Maya, Anelizze Castro-Martínez, Marisol Chacón, Willi Cheung, Evelyn S. Crescini, Peter De Hoff, Emily Eisner, Sawyer Farmer, Abbas Hakim, Laura Kohn, Alma L. Lastrella, Elijah S. Lawrence, Sydney C. Morgan, Toan T. Ngo, Alhakam Nouri, R Tyler Ostrander, Ashley Plascencia, Christopher A. Ruiz, Shashank Sathe, Phoebe Seaver, Tara Shwartz, Elizabeth W. Smoot, Thomas Valles, Gene W. Yeo, Louise C. Laurent, Rebecca Fielding-Miller, Rob Knight

## Abstract

Monitoring severe acute respiratory syndrome coronavirus 2 (SARS-CoV-2) on surfaces is emerging as an important tool for identifying past exposure to individuals shedding viral RNA. Our past work has demonstrated that SARS-CoV-2 reverse transcription-quantitative PCR (RT-qPCR) signals from surfaces can identify when infected individuals have touched surfaces such as Halloween candy, and when they have been present in hospital rooms or schools. However, the sensitivity and specificity of surface sampling as a method for detecting the presence of a SARS-CoV-2 positive individual, as well as guidance about where to sample, has not been established. To address these questions, and to test whether our past observations linking SARS-CoV-2 abundance to *Rothia* spp. in hospitals also hold in a residential setting, we performed detailed spatial sampling of three isolation housing units, assessing each sample for SARS-CoV-2 abundance by RT-qPCR, linking the results to 16S rRNA gene amplicon sequences to assess the bacterial community at each location and to the Cq value of the contemporaneous clinical test. Our results show that the highest SARS-CoV-2 load in this setting is on touched surfaces such as light switches and faucets, but detectable signal is present in many non-touched surfaces that may be more relevant in settings such as schools where mask wearing is enforced. As in past studies, the bacterial community predicts which samples are positive for SARS-CoV-2, with *Rothia* sp. showing a positive association.

**Importance:** Surface sampling for detecting SARS-CoV-2, the virus that causes coronavirus disease 2019 (COVID-19), is increasingly being used to locate infected individuals. We tested which indoor surfaces had high versus low viral loads by collecting 381 samples from three residential units where infected individuals resided, and interpreted the results in terms of whether SARS-CoV-2 was likely transmitted directly (e.g. touching a light switch) or indirectly (e.g. by droplets or aerosols settling). We found highest loads where the subject touched the surface directly, although enough virus was detected on indirectly contacted surfaces to make such locations useful for sampling (e.g. in schools, where students do not touch the light switches and also wear masks so they have no opportunity to touch their face and then the object). We also documented links between the bacteria present in a sample and the SARS-CoV-2 virus, consistent with earlier studies.

## Body

Environmental monitoring for severe acute respiratory syndrome coronavirus 2 (SARS-CoV-2) RNA by reverse transcription-quantitative polymerase chain reaction (RT-qPCR) is increasingly gaining acceptance. In the Safer at School Early Alert (SASEA) (https://saseasystem.org/) project, daily surface swabbing was employed as part of an effort to detect coronavirus disease 2019 (COVID-19) cases in nine elementary schools. This study identified 89 clinically positive COVID-19 cases, 33% preceded by a room-matched surface positive (1). As pandemic response measures like SASEA become more widely implemented, understanding where SARS-CoV-2 signatures will most likely be found reduces cost and labor of surface swabbing in large facilities. Previous work has focused on sampling arbitrary surfaces in isolation and congregate care facilities, homes, and hospitals, with varying detection performance obscuring which surfaces are best for monitoring COVID-19 spread (2-6). Counterintuitively, high-touch hospital surfaces expected to accumulate viral load, including door handles and patient bed rails, can yield *lower* SARS-CoV-2 detection rates, presumably because they are cleaned more often (7-8).

Most microbes in the built environment come from human inhabitants (9-11). Oral, gut, and skin microbiomes of COVID-19 patients change during disease (8,12-13); therefore, SARS-CoV-2 positive built environmental samples may differ in *bacterial* communities from SARS-CoV-2 negative samples. This has been documented in a hospital setting, with associations between SARS-CoV-2 status (Detected/Not Detected) and both overall microbial community and *Rothia* spp. specifically (8).

To extend these results to a residential setting and understand how SARS-CoV-2 is distributed in the living space of an infected individual, we performed environmental sampling in the apartments of three people who recently tested positive for COVID-19 (Sup. Fig. S1) while quarantined in an isolation facility. On the day of swabbing, each quarantining individual provided an anterior nares swab sample (Average Cq: 29.5, 28.4, 28.6 for Apartments A, B, and C respectively). Although apartments differed in size, floor plan, and features (furniture, appliances, etc.), similar features at similar densities were swabbed across all three (n=140,116,125).

Each sampled surface was swabbed twice in immediately adjacent locations: first with a swab premoistened and stored in 95% ethanol, then by a second swab premoistened and stored in a 0.5% SDS w/v solution (Supplementary Methods). Ethanol samples underwent 16S V4 rRNA gene amplicon (16S) sequencing, and SDS samples underwent RT-qPCR for SARS-CoV-2 detection. 16S sequences were demultiplexed, quality filtered, and denoised with Deblur (14) in Qiita (15) using default parameters. Resulting feature tables were processed using QIIME2 (16).

### Findings

We collected 381 matched 16S and SARS-CoV-2 surface samples from the three apartments, of which 178 (47%) were positive for SARS-CoV-2 (Fig 1) (Table 1). Apartments A and C had comparable positivity rates (53% and 61%, respectively), but Apartment B was substantially lower (24%). In all three apartments, the rate of detection was highest in the bedroom (72% on average vs 47% overall). We estimated surface viral load, in viral Genomic Equivalents (GE’s), from Cq’s using published regression curves (17) and mapped resulting viral loads onto 3D renderings of each apartment. High-touch surfaces, including handles and switches, had highest viral load across all apartments, followed by floor samples and then high-use objects (fridge, sinks, toilets, beds) (Fig. 1). The maps for each apartment were studied to understand patterns of SARS-CoV-2 detection and deposition by room use. In the kitchens, objects with planar faces and handles, such as the refrigerator, cabinets, and drawers, revealed that only the touched handles had detectable RT-qPCR signal (Fig. 1C inset, as an example). We could not detect viral RNA on adjacent planar faces, which were presumably breathed on but not touched.

**Figure 1.**
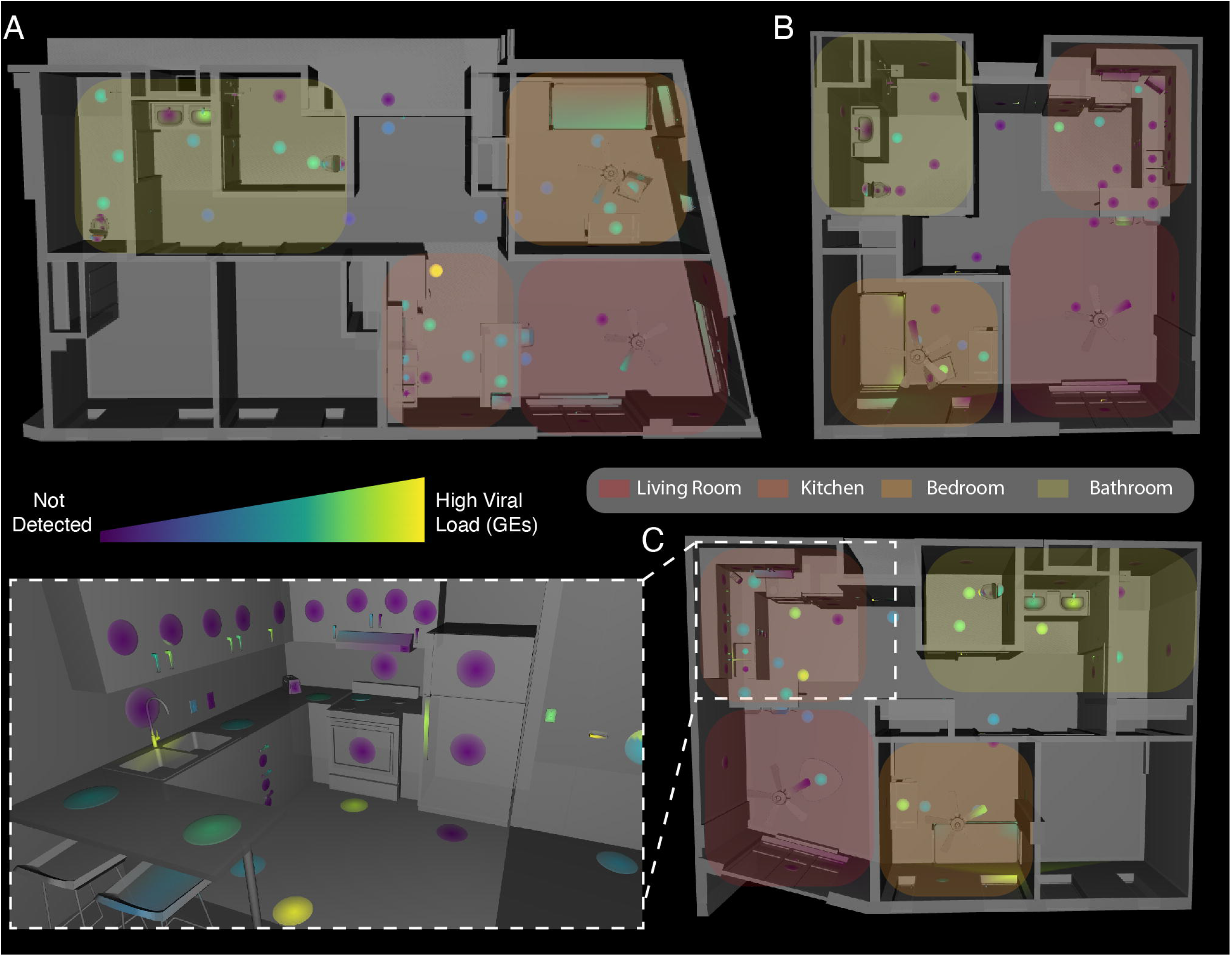
Distribution of SARS-CoV-2 viral load in isolation dorm apartments. (A-C) Floor plans for each apartment highlighting where SARS-CoV-2 RNA signatures were detected. (Inset) 3D rendering of the kitchen in Apartment C showing SARS-CoV-2 viral load in Genomic Equivalents (GEs) mapped to features in that room.

For quality control of 16S sequencing from low-biomass samples, we sequenced surface swabs from the apartments together with positive and negative controls using KatharoSeq (Supplementary Methods) (Sup. Fig. S2A) (18). Of 381 samples that underwent 16S sequencing, 121 fell below the KatharoSeq threshold and were excluded (Sup. Fig. 2C). Informed by alpha rarefaction curves (Sup Fig 2B), remaining samples were rarefied to 4000 features, removing an additional 36 samples from the analysis. Therefore, 157 samples were excluded from downstream analyses (122 SARS-CoV-2 negative matched swabs, 35 positive) (Sup Fig 2C).

Bacterial alpha diversity analysis demonstrated that 16S amplicon read count associated with SARS-CoV-2 detection status (Sup. Fig. S3). Forward stepwise redundancy analysis (RDA) using the unweighted UniFrac beta diversity metric identified four non-redundant variables of significant effect size (apartment, surface type, type of room, and SARS-CoV-2 detection status) which accounted for 45.4% of the variation in the data (Sup. Fig. 4B). Analyzed by apartment, only in apartment B did virus detection lack significant effect. When subsetting the entire dataset by room type, detection status had a significant effect on variability across all rooms.

To test whether the bacterial community predicted SARS-CoV-2 status, we built a random forest classifier using sOTU data. The overall Area Under the Precision-Recall Curve (AUPRC) was 0.78, suggesting a statistically significant association, but insufficiently strong to predict SARS-CoV-2 status of a single sample from the bacterial community (Fig. 2A). Cross-application of models trained from one apartment or room type to other apartments or room types generally performed well (AUPRC=0.7-0.96), suggesting generalizable associations (Fig. 2B). We also applied multinomial regression to our dataset to identify differentially abundant microbes between SARS-CoV-2 status groups. The top 32 features identified by the random forest classifier and the ranked log-fold-changes in feature abundance from the multinomial regression are shown in Figure 2C. Agreeing with previously published findings, *Rothia dentocariosa* was one of the top features identified by the classifier and was relatively positively associated with SARS-CoV-2 positive samples in the regression (8,12). Six sOTUS belonging to members of the genus *Corynebacterium* were also highly ranked as predictive for positive samples.

**Figure 2.**
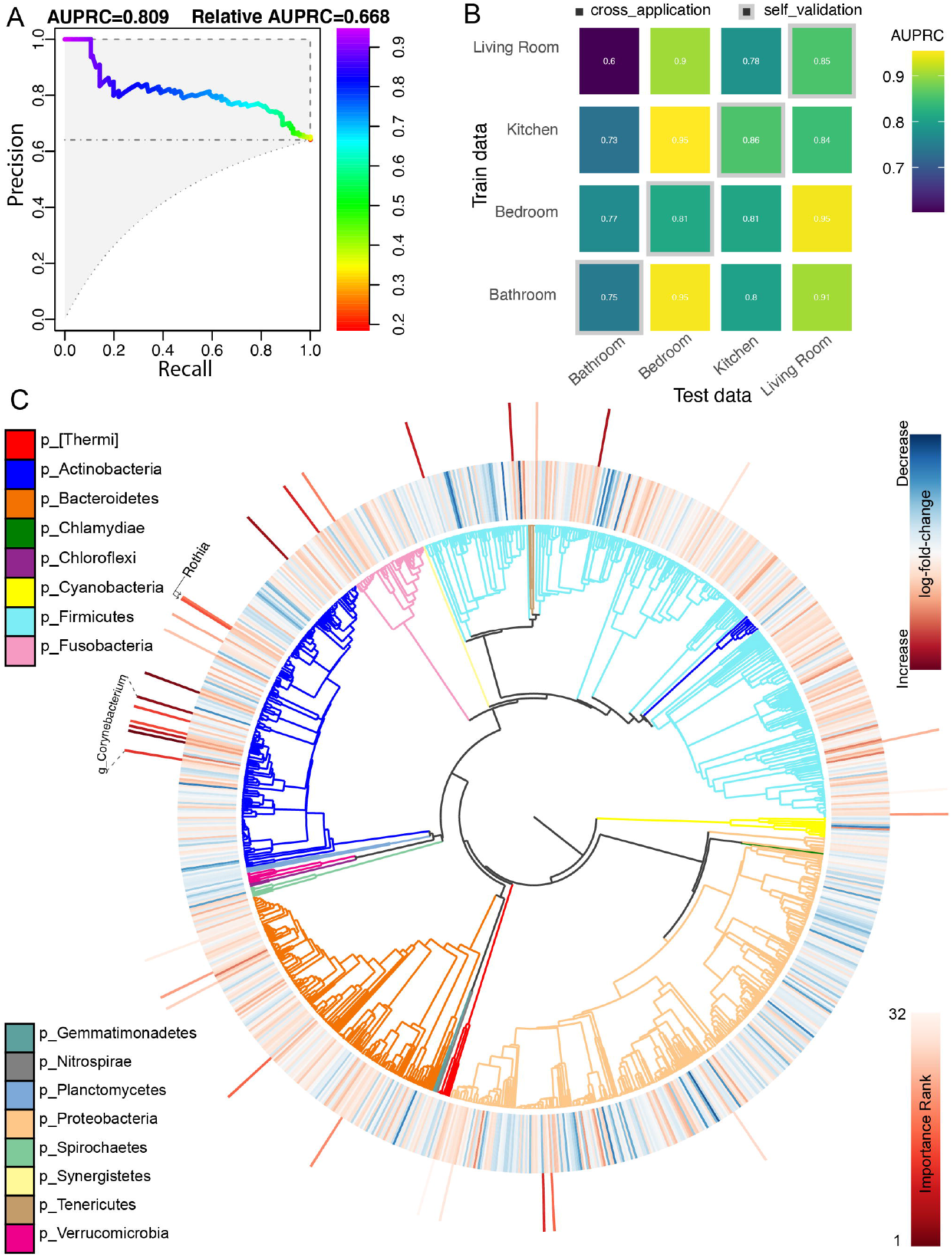
(A) Area under the precision-recall curve showing the overall prediction performance of the random forest classifiers when trained on the features from two apartments and cross validated on the remaining apartment. (B) Confusion matrix showing per-room type classifiers when cross-applied on the remaining room types. The diagonal represents self validation. (C) Phylogenetic tree visualization (EMPress) where the differentially-abundant features between SARS-CoV-2 status groups identified by multinomial regression (Songbird) are plotted on the inner ring, and the ranked sOTUs identified as important by the random forest classifier are indicated on the outer ring.

## Discussion

Our results show that detailed spatial mapping of SARS-CoV-2 RNA abundance and associated bacterial signatures from built environment surfaces provides useful insight into potential sampling locations and associations between the viral and bacterial components of the microbiome. In the residential setting, high-touch surfaces have especially high viral loads, although confirming this with detailed spatial maps in other settings (hospitals, isolation hotels, schools) may be useful for guiding sampling designs. We note that sensitivity of arbitrary single surface sampling to detect presence of even an unmasked SARS-CoV-2 individual is low, so multiple samples or samples from selected surfaces should be collected. These results reinforce the utility of surface sampling as a cost-effective method for locating SARS-CoV-2 signals in the environment.

Our findings also corroborate SARS-CoV-2 associated changes in the microbiome published previously. *Rothia dentocariosa* specifically has been identified across different sample types in diverse settings, although reasons for these associations remain unclear. We also see multiple sOTUs belonging to the genus Corynebacterium predictive of a SARS-CoV-2 detection event, in contrast to the findings of another study that found Corynebacterium significantly decreased in the oral microbiome of individuals with COVID-19 (12). We hypothesize that Corynebacterium signal in this study might be evidence of human skin contamination of indoor surfaces through contact, leading to SARS-CoV-2 deposition on surfaces. It has been established that the occupants of a room contribute to the environmental microbiota, but our findings are among the first to demonstrate that disease-associated changes in the microbiome are mirrored in the built environment.

## Data Availability

All sequencing data produced are available online at https://qiita.ucsd.edu/study/description/13957

https://qiita.ucsd.edu/study/description/13957

## Acknowledgements

This research was supported by NIH grant (K01MH112436) to RFM, and the County of San Diego Health and Human Services Agency (Contract 563236). We thank Min Yi Wu, Bing Xia, Daniel Maunder, Michal Machnicki, Bhavika K. Kapadia, and Lizbeth Franco Vargas for their support with environmental SARS-CoV-2 detection as part of the EXCITE Lab.

## Figure Legends

**Supplementary Figure 1.**
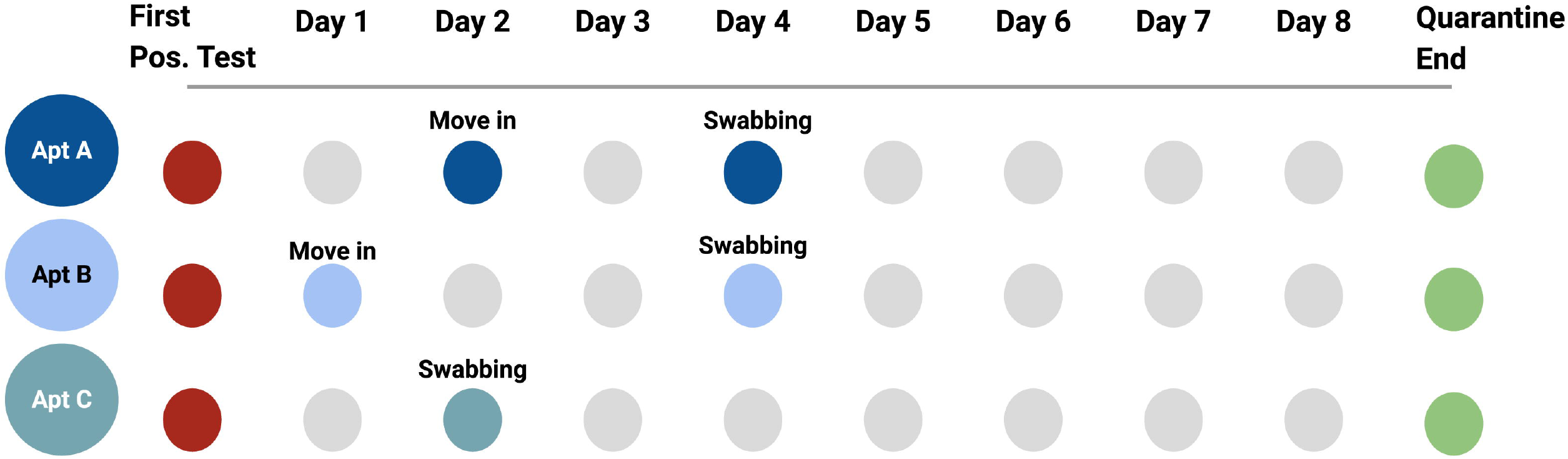
Timeline of events from first positive test to the end of the individual’s quarantine period. Apartment C has no move in date because the individual quarantined in place.

*Supplementary Table 1. Environmental samples with detectable SARS-CoV-2 per apartment and room type*.

**Supplementary Figure 2.**
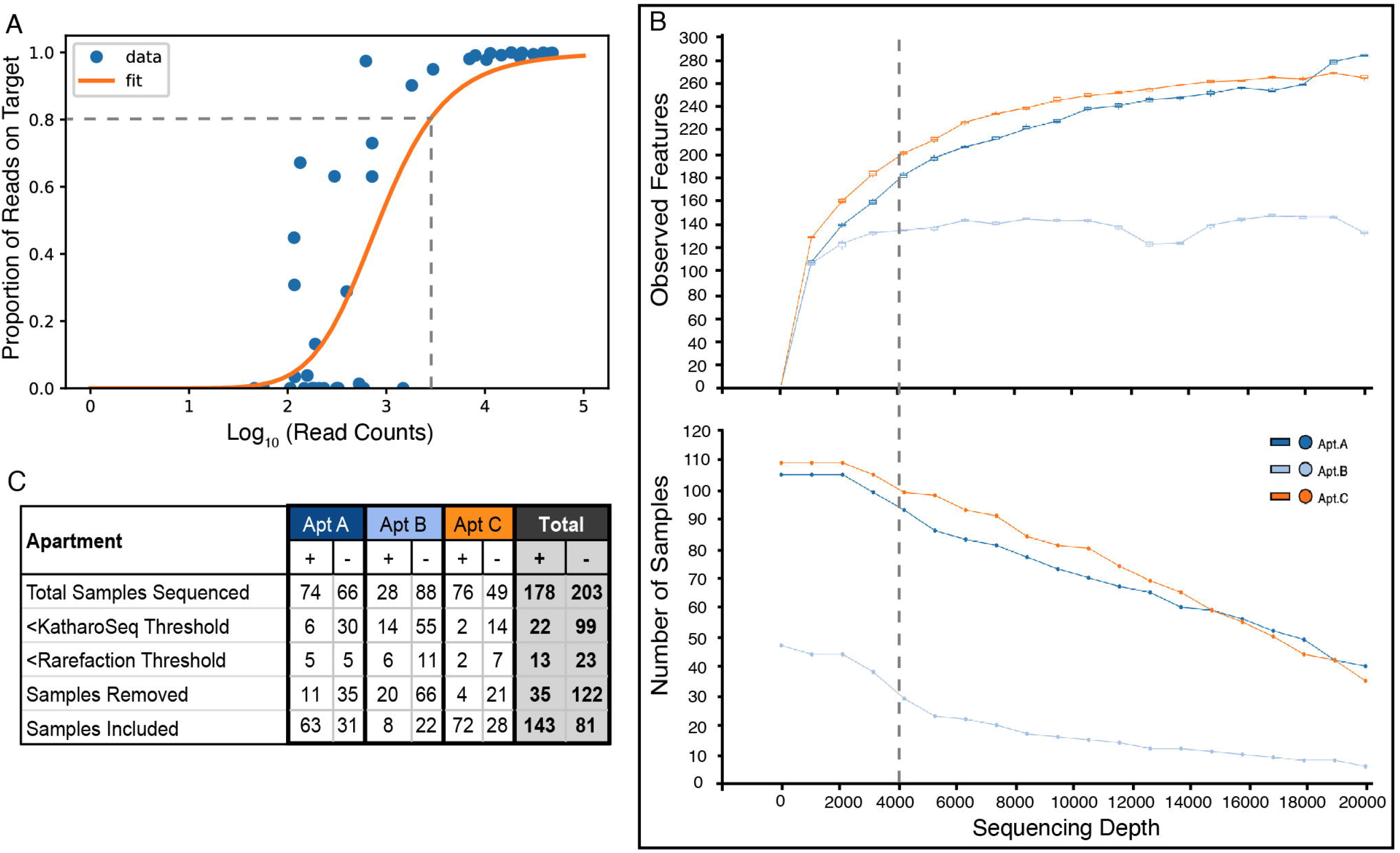
Exclusion criteria for low biomass samples. (A) Diluted stock of a KatharoSeq positive control was sequenced along with the environmental samples and the resultant reads underwent pre-processing as detailed in the Supplementary Methods.The KatharoSeq Threshold (dashed lined), a minimum number of reads derived from a fitted allosteric sigmoidal curve, corresponds to a sequencing depth where at least 80% of the positive control reads are taxonomically classified to the appropriate target organisms (B) Top panel: Rarefaction curve showing observed features (alpha diversity metric) as a function of sequencing depth. Bottom panel: Graph showing how many samples would be included in downstream analysis as a function of rarefaction depth. (C) Table showing how many samples were removed at the KatharoSeq and Rarefaction thresholds and overall.

**Supplementary Figure 3.**
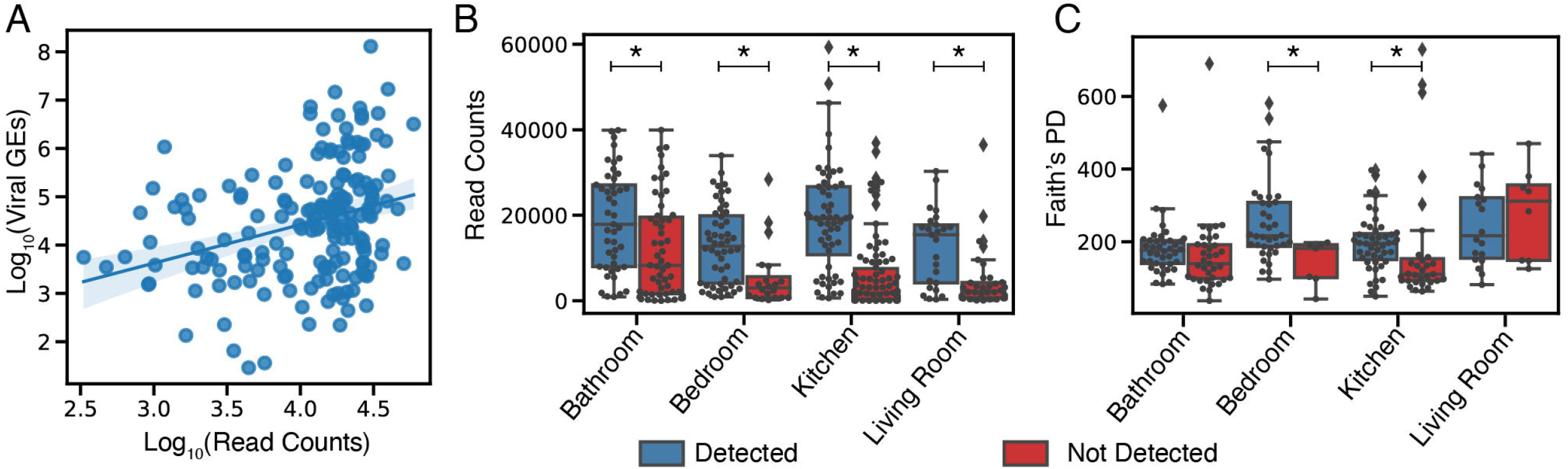
Correlation between microbial biomass/diversity and SARS-CoV-2 detection. (A) Number of 16S reads in SARS-CoV-2 positive samples shows significant correlation with SARS-CoV-2 viral load (GE’s) (*Pearson correlation, r=0.3, p=3×10^−5^*). (B) Read counts are significantly different between positive and negative samples when compared within room types (Mann-Whitney U tests, p≤0.003). (C) Alpha diversity (Faith’s PD) shows a weaker significance between positive and negative samples when compared within room types with only the bedroom and kitchen showing a significant difference between positive and negative samples (Mann-Whitney U tests, p=0.01).

**Supplementary Figure 4.**
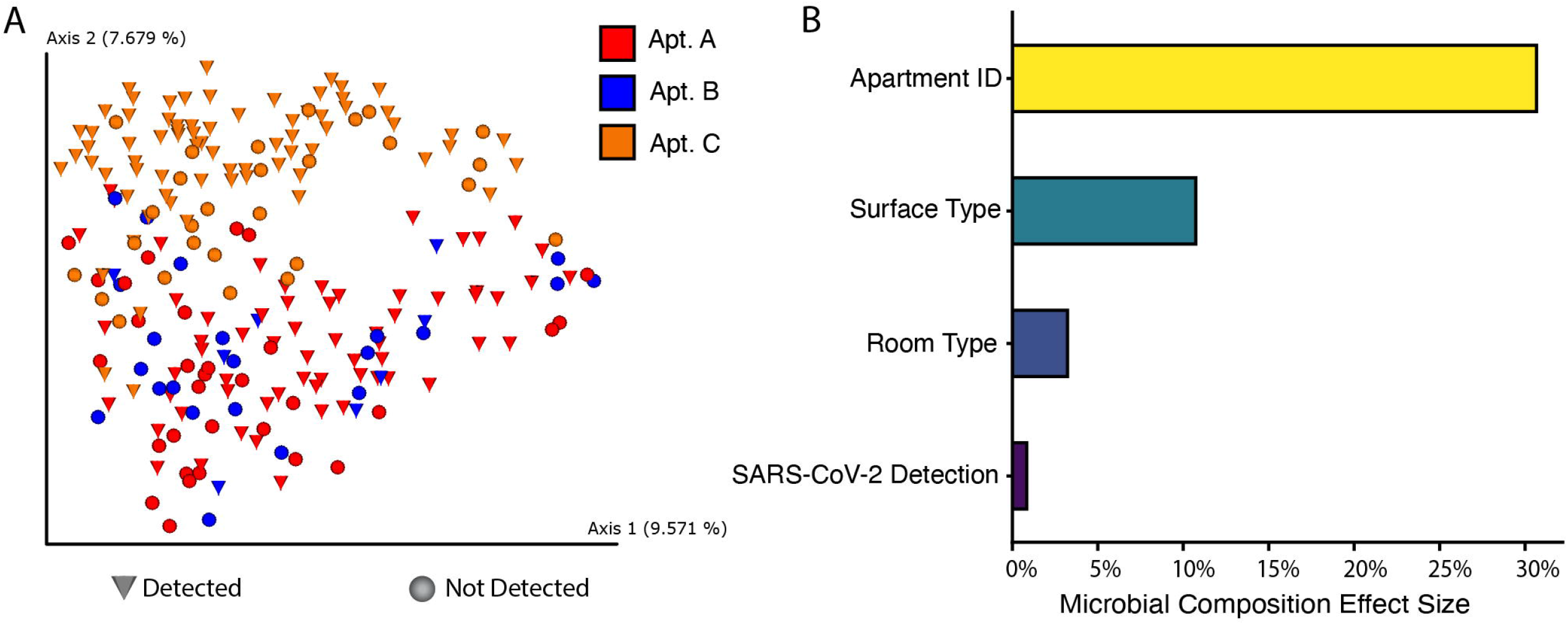
Beta diversity analysis identifies the factors that contribute most to the separation of the data. (A) Principal coordinates analysis of the Unweighted Unifrac distance matrix shows that a major driver in the separation of this data is which apartment the samples came from. (B) Barplot showing the statistically significant effect sizes for non-redundant variables returned by RDA analysis. The largest effect size was explained by apartment (30.7%, p=0.0002), followed by surface material type (10.7%, p=0.0002), room type (3.2%, p=0.0004), and SARS-CoV-2 detection status (0.84%, p=0.01).

## Notes

### Competing Interest Statement

The authors have declared no competing interest.

### Author Declarations

Patients were recruited to the study via phone call, enrolled into an IRB-approved study (UCSD protocol 200477), and confirmed to be active UCSD students isolating in the isolation dorms.

