## Supplementary Materials and Methods for "SARS-CoV-2 Distribution in Residential Housing Suggests Contact Deposition and Correlates with *Rothia* sp"

*Patient Recruitment:*

In an effort to identify patients isolating in similar residential settings, the patient population focused on University California San Diego (UCSD) students isolating in the established, on-campus UCSD isolation dorm housing. Cases were identified through the UCSD Health system as COVID-19 positive outpatients with a positive anterior nares clinical RT-qPCR assay from the UCSD EXCITE (EXpedited COVID-19 IdenTification Environment) laboratory. Patients were recruited to the study via phone call, enrolled into an IRB-approved study (UCSD protocol 200477), and confirmed to be active UCSD students isolating in the isolation dorms. Three students were enrolled in the study: two of the students isolated in the on-campus isolation dorms, and the third isolated in their on-campus residence (graduate housing with similar architecture and design as the on-campus isolation dorms).

*Surface Swabbing:*

For each paired sample, two 1 mL sample collection tubes (ThermoFisher Scientific, 3740TS) were prepared. One tube contained 800 µL of 0.5% w/v sodium dodecyl sulfate (SDS) (Acros Organics, 230420025) in water and was used for detecting SARS-CoV-2, and the second tube contained 95% spectrophotometric-grade ethanol solution (Sigma-Aldrich #493511) which was designated for 16S sequencing. To recover genetic material from the surfaces, a prewashed cotton swab (Puritan, 806-WC) was pre-moistened with the ethanol solution and then used to vigorously swab the surface. The cotton end of the swab was then placed back into the sample collection tube and broken at the designated break point. The process was then immediately repeated on an adjacent site of the same surface with a flocculated tip swab (Affordable IHC Solutions) pre-moistened with the SDS solution, minimizing overlap between swabbed areas.

*Viral Nucleic Acid Extraction and RT-qPCR:*

Swabs stored in SDS were subjected to SARS-CoV-2 RT-qPCR detection following methods previously described (1). Briefly, 150µL of the SDS solution were extracted with Omega MagBind Viral DNA/RNA kit (Omega Bio-Tek, M6246) on Kingfisher Flex (ThermoFisher Scientific) instruments. Viral gene detection was performed using a miniaturized TaqPath™ COVID-19 Combo Kit (ThermoFisher Scientific, A47814) assay on a QuantStudio 7 Pro with a 384-well sample block (ThermoFisher Scientific).

*Microbial Nucleic Acid Extraction:*

Sample plating and extractions of all surface swabs were carried out in a biosafety cabinet Class II in a BSL2+ facility. Cotton tipped swabs suspended in 95% ethanol were plated into bead plates from 96 MagMAX™ Microbiome Ultra Nucleic Acid Isolation Kits (A42357 Thermo Fisher Scientific, USA). Following the KatharoSeq low biomass protocol (2), each sample processing plate included eight positive controls consisting of 10-fold serial dilutions of a microbial standard consisting of a gram negative *Paracoccus spp.* and gram positive *Bacillus subtilis* ranging from 5 to 50 million cells per extraction, and 3 negative controls (Blanks, sample-free lysis buffer). Nucleic acid extraction and purification was performed following methods previously described (3). Briefly, samples were extracted in plates using the

MagMAX$^{TM}$ Microbiome Ultra Nucleic Acid Isolation Kit (Applied Biosystems$^{TM}$), following manufacturer specifications, in KingFisher Flex$^{TM}$ robots (Thermo Fisher Scientific, USA), including a bead beating step in a TissueLyser II (Qiagen, Germany) at 30 Hz for 2 min.

*16S Sequencing:*

16S rRNA gene amplification was performed according to the Earth Microbiome Project protocol (4). Briefly, the V4 region of the 16S rRNA gene was targeted for amplification in a miniaturized reaction (5) using the 515f-806r primers with Golay error-correcting barcodes. Amplicons were pooled at equal volumes and the pool was purified with a QIAquick PCR purification kit (QIAGEN). The pooled libraries were sequenced on a MiSeq (Illumina) instrument with a MiSeq Reagent 300 cycle v2 Kit, with the appropriate sequencing primers.

*Estimating genomic equivalents and microbial biomass:*

To estimate viral genomic equivalents for each sample, we used published standard curves relating average Cqs from RT-qPCR to known SARS-CoV-2 viral particle concentrations (in GE's from digital droplet PCR) used to inoculate a variety of indoor surfaces (1). The equation used depended on which qualitative category the surface materials belonged to: rough (carpet, fabric) or smooth (e.g., acrylic, steel, glass, ceramic tile). The relationship between Cqs and GEs for rough materials is [GEs = -0.52 x (Avg Cq) + 39.90] while for smooth materials the equation used was [GEs = -0.77 x (Avg Cq) + 40.41].

We used an equivolumetric sequencing library pooling approach which allowed us to correlate biomass to 16S amplicon counts (6).

*Data processing:*

16S sequences were demultiplexed, quality filtered, and denoised with Deblur (7) in Qiita (8) using default parameters. Resulting feature tables were processed using QIIME2 (9). Sequencing data available in Qiita study ID: 13957.

*Katharoseq***:**

In addition to the 381 samples that underwent 16S sequencing, three negative controls (blanks) and eight positive controls (a serially diluted bacterial stock, see *Microbial Nucleic Acid Extraction*) were included in each 96-well extraction plate. The positive controls were used to determine the threshold read count for which at least 80% of sequencing reads align to the positive controls (10).

*Alpha Diversity:*

To explore the relationship between microbial biomass and SARS-CoV-2 status, we compared the estimated SARS-CoV-2 viral load in GEs and the number of raw 16S reads for all samples. The Pearson correlation coefficient was calculated to determine if the two measurements had a linear relationship [log(16S Read Counts), log(GE's)]. The relationship between biomass (16S read count) and SARS-CoV-2 detection status (Detected/Not Detected) for samples in the same room type was tested with a Kruskal-Wallis H test. For the stringently filtered feature tables, differences in Faith's Phylogenetic Diversity (Faith's PD) between SARS-CoV-2 detection status within each room were also tested using a Kruskal-Wallis H test. 2D Figures were made using matplotlib (11).

*Beta Diversity:*

We used the unweighted Unifrac phylogenetic distance (12-13) to explore how the microbial samples compare to each other. To quantify the effect size of different categorical variables on our data, redundancy analysis (RDA) was applied to the unweighted Unifrac principal coordinates. RDA estimates the contributions of individual and combined effects of multiple covariates using the *varpart* function in R to perform linear constrained ordination (14). 2D Figures were made using matplotlib (11) and EMPeror (15).

*Differential Abundance:*

To prepare the data for differential abundance we filtered the unrarefied feature table to exclude features present in fewer than 10 samples and samples with depth less than 1000. This resulted in a table of 258 samples and 1047 sOTUs. We performed multinomial regression using Songbird (16) accounting for viral detection status, apartment, surface type, and indoor space classifier as covariates. We used 5000 epochs and a learning rate of 0.0001 as hyperparameters. Additionally, we specified a 3:1 split of training:testing samples for cross validation. To ensure that our model was not overfitting we fit a null regression model with no covariates using the same hyperparameters. Comparing the two models we found a positive pseudo-$Q^2$ value of 0.059, indicating that our regression model outperformed the null model.

*Random Forest Classifier:*

We performed machine learning analysis on the bacterial portion of the built environment surface microbiome from 16S sequencing to predict the samples' SARS-CoV-2 status from paired RT-qPCR detection results. Random forest classifiers were trained and tested following a leave-one-site-out-cross-validation (LOSOCV) approach: the classifier was trained with samples from N-1 sites and its performance was tested in the remaining site using a precision-recall curve (Area Under the Precision Recall Curve (AUPRC), and Relative AUPRC). Classifiers were trained on sOTU-level features with tuned hyperparameters as 20-time repeated, LOSOCV, with sites resolved at the apartment_id (Fig. 2A) and room_type (Fig. 2B) levels using the R caret package(17). The classifiers' performance was evaluated with AUPRC based on the samples' SARS-CoV-2 status predictions of the holdout test site using the R PRROC package (18). The importance of each sOTU for the prediction performance of the classifiers was estimated by the built-in random forest scores in a 100-fold cross validation. We ranked the top 32 important features by their average ranking of importance scores across the 100 classification models. Relevant codebase for machine learning analysis is available at https://github.com/shihuang047/crossRanger and is based on random forest implementation from R ranger package (19).

*Phylogenetic Tree visualization:*

To identify phylogenetic clades important for the prediction of SARS-CoV-2 status from environmental surface samples we visualized the top 32 important features identified by the random forest classifier and the ranked differentially abundant features between SARS-CoV-2 status groups  from multinomial regression using EMPress (20).

*3D Mapping:*

3D models were provided by UC San Diego's Housing, Dining, and Hospitality department. A circular target was placed on all swabbed locations in each apartment. 3D coordinates were picked following published methods (ref) (https://github.com/MolecularCartography/ili), and merged with viral load (in GEs) data for visualization. 3D models and merged data (coordinates and viral load) were visualized in ili (21).
